# Tenofovir-DF versus Hydroxychloroquine in the Treatment of Hospitalized Patients with COVID-19: An Observational Study (TEDHICOV)

**DOI:** 10.1101/2021.03.24.21252635

**Authors:** Mario Cornejo-Giraldo, Nelson Rosado, Jesús Salinas, Nelson Aspilcueta, Eduardo Bernales, Jimmy Lipa, Johanna Coacalla, Yoisi Flores, Pamela Leon, Claudia Chamby

## Abstract

**Background:** Although several therapeutic agents have been suggested for the treatment of the disease caused by the Coronavirus of the year 2019 (COVID-19), no antiviral has yet demonstrated consistent efficacy.

**Methods:** The results of an observational study comparing Tenofovir-DF (TDF) with Hydroxychloroquine (HCQ) in the treatment of hospitalized patients with COVID-19 with evidence of pulmonary compromise and the vast majority with supplemental oxygen requirement are presented. Patients received HCQ consecutively at the dose of 400 mg. 12 hourly for 01 day and then 200 mg. every 8 to 12 hours PO for 5 to10 days; or TDF 300 mg. per day PO for 7 to 10 days. The primary outcomes of the study were the differences between the two groups regarding: hospital stay, the need for intensive care or mechanical ventilation (ICU / MV) and mortality.

**Results:** 104 patients were included: 36 in the HCQ group and 68 in the TDF group. The unadjusted primary outcomes were: LOS (length of stay) 16.6 ± 12.1 for HCQ versus 12.2 ± 7.0 days for TDF (p = o.o102); need for admission to ICU / mechanical ventilation (MV): 61.1% for HCQ versus 11.8% for TDF (p = o.ooo); and mortality: 50.0% for HCQ and 8.8% for TDF (p = o.ooo). The patients in the HCQ group had significant differences at admission compared to those in the TDF group regarding: male sex, cardiovascular risk factor, greater respiratory involvement and higher glucose and creatinine levels, lower albumin levels and higher. Inflammatory markers. When the outcomes were adjusted for these baseline differences, in the multiple regression model for LOS, it was found that TDF decreased the hospital stay by 6.10 days (C.I.: −11.97 to −2.40, p = o.o42); In the logistic regression model for the need for ICU / MV, it was found that the use of TDF had an O.R. of 0.15 (C.I.: 0.03-0.76, p = o.o22); and for the Cox proportional hazards model for mortality, the H.R. was 0.16 for TDF (C.I.: 0.03-0.96, p = o.o41). In the estimation model of the treatment effects by regression adjustment, it was found that TDF decreased the stay by −6.38 days (C.I.: −12.34 to −0.42, p = o.o36); the need for ICU / MV at −41.74% (C.I.: −63.72 to −19.7, p = o.ooo); and mortality by −35.22% (C.I.: −56.47 to −13.96, p = o.oo1).

**Conclusion:** TDF may be an effective antiviral in the treatment of COVID-19. Some of its advantages include: its wide availability, cost and oral presentation. Randomized clinical trials are imperatively required to confirm this possibility.

## TEXTO

Since its appearance in Wuhan-China in December 2019 [1], COVID-19 caused by Coronavirus 2 associated with Severe Acute Respiratory Syndrome (SARS-CoV-2) has spread rapidly throughout the world. In Peru, the Pandemic arrived in March 2020, first in Lima, the capital of Peru, and that same month it reached Arequipa. Until the first days of December, COVID-19 cases in Peru have reached about 970,000 confirmed cases with at least 30,000 deaths officially reported [2]; in Arequipa we reached about 145,000 cases with 2,305 deaths for that same date [3]. Arequipa is one of the largest cities in Peru, located about 2,300 meters above sea level and has a population of about one million inhabitants. The Carlos Alberto Seguín Escobedo National Hospital (CASE) is a complex hospital and the head of the southern care network of the social security (EsSalud) with an attached population of about 700,000 people. Since the beginning of the outbreak in the city, it was designated as a Hospital for COVID patients where most of the insured patients and with more severe disease were treated.

In general, the accepted management of COVID-19 is primarily symptomatic and oxygen support and ventilatory support when required. Dexamethasone has been shown to be effective in hospitalized patients with a requirement for supplemental oxygen [1-3]. Regarding agents with direct action against the virus, initial hopes focused on remdesivir, Hydroxychloroquine, lopinavir / ritonavir and interferon-β. Of these agents, only remdesivir appears to have any favorable action against COVID-19 [4-10]. In contrast, various studies have shown the lack of efficacy of the other schemes [11-20]. We present here an observational study, the first one reported to our knowledge, of the use of tenofovir-DF (disoproxil fumarate) (TDF) compared with Hydroxychloroquine in hospitalized patients, the vast majority of them with moderate or severe disease caused by COVID-19 with supplemental oxygen requirement, in categories 5 to 9 of the WHO clinical progression scale [21].

## Methods

All hospitalized patients with COVID-19 from March 2020 to May 30, 2020 at the CASE-EsSalud National Hospital with a PCR-confirmed diagnosis of SARS-CoV-2 were included. All patients gave informed consent. The first patients received Hydroxychloroquine (HCQ) with lopinavir / ritonavir (LPV / r) or azithromycin (AZM) which were the national standard of therapy at that time [1]. Tenofovir disoproxil fumarate (TDF) was started as monotherapy in late April.

Inclusion criteria were: hospitalized adults over 18 years of age with a positive molecular test for SARS-CoV-2, computed tomography (TC scan) compatible with “COVID pneumonia” and / or in need of supplemental oxygen, who had received HCQ or TDF as therapy, that the duration of treatment had been at least three days and under informed consent. The exclusion criteria were: negative molecular test for SARS-CoV-2, no hospitalization, negative CT scan for “COVID pneumonia” and / or no supplemental oxygen requirement for COVID-19, who had not received either HCQ or TDF or both drugs at the same time, who had received HCQ or TDF for two days or less and had no informed consent.

The standard treatment with Hydroxychloroquine (HCQ) was: 400 mg every 12 hours PO for 01 days and then 200 mg every 8-12 hours for 5 to 10 days; azithromycin (AZM) 500 mg PO the first day and then 250 mg once daily PO for an additional 2 to 4 days; lopinavir / ritonavir (LPV / r) 400/100 mg every 12 hours PO for 7-10 days. Regarding treatment with tenofovir, monotherapy with tenofovir-DF (disoproxil fumarate) (TDF) was used at the usual dose of 300 mg PO per day for 7 to 10 days. In addition, all patients received usual symptomatic and supportive treatment.

The primary outcomes were mortality, the need for admission to the Intensive Care Unit (ICU) and / or Mechanical Ventilation, and LOS (length of stay) in both groups. Furthermore, demographic factors, risk factors, vital / respiratory functions, and laboratory results of both groups were compared. The tests closest to the time of admission to the hospital were taken into consideration.

It was decided to assess the severity of pulmonary involvement mainly by means of SaFi (oxygen saturation by pulse oximetry divided by the fraction of inspired oxygen) as it is a quantitative and more accurate variable than ordinal characterization.

### Statistical Analysis

quantitative variables are expressed as mean ± standard deviation, qualitative variables are expressed as percentages (%). In the univariate analysis, the Student’s t test was used for the means and the chi2 test for nominal or ordinal variables. Kaplan-Meier survival curves were evaluated 30 days after admission to the hospital to assess mortality. The multivariate analysis of the variables that were significant in the univariate analysis was carried out by the Cox regression model in the evaluation of mortality; the multivariate linear regression model for the evaluation of the stay and the “log rank” model for the evaluation of admission to the Intensive Care Unit (ICU) or the need for mechanical ventilation. In addition, models were developed to estimate the effects of TDF treatment by regression adjustment for the primary outcomes of this observational study: hospital stay, need for ICU (intensive care unit) and / or MV (mechanical ventilation) and mortality. The Stata® software version 14 was used and p <o.o5 was considered significant.

## Results

In the study period, there were 162 hospitalized patients diagnosed with COVID-19, of which 58 patients were excluded for the following reasons: they had received 2 days or less of therapy (23), or had not received either of the two schemes (21), or they were PCR negative for SARS-CoV-2 (7), or because they had received both HCQ and TDF (7). Thus, 104 hospitalized patients with a diagnosis of COVID-19 confirmed with PCR, compatible CT scan and / or with supplemental oxygen requirement were included. Of these, 36 received HCQ and 68 TDF.

The demographic variables and risk factors are shown in Table No. 1. In the univariate analysis, a statistically significant difference was only found with regard to a higher frequency of males in the HCQ group vs. TDF: 75.0% vs. 54.4% (p = o.o370), respectively; and in the presence of a cardiovascular risk factor 44.4% vs. 14.7% (p = o.oo1), respectively. This cardiovascular risk factor was arterial hypertension in all cases. The mean age of the patients in the HCQ group was 60.5 ± 14.5 years (range 32 to 89 years) and that of the TDF group was 55.8 ± 15.8 years (range 28 to 87 years): although the patients in the HCQ group they were about 5 years older than those in the TDF group, there was no statistically significant difference (p = o.o705). The mean time of illness upon admission to hospital was 6.9 ± 2.0 days (range 3 to 12 days) in the HCQ group, while in the TDF group it was 7.6 days ± 2.6 days (range 2 to 14 days). days) without statistically significant difference (p = o.103).

**Table No. 1.** Tenofovir-DF versus Hydroxychloroquine: comparative demographic variables between the two groups.

| Variables | Hydroxychloroquine<br>36 patients | Tenofovir<br>68 patients | p |
| --- | --- | --- | --- |
| Age (years ± S.D.) | 60.5 ± 14.5 | 55.8 ± 15.8 | 0.0705 |
| Sex Male (No. %) | 27/36 (75.0 %) | 37/68 (54.4 %) | 0.0370 |
| Sick time (days ± S.D.) | 6.9 ± 2.0 | 7.6 ± 2.6 | 0.1003 |
| Risk factor (No. %) | 22/36 (61.1%) | 28/68 (41.2%) | 0.052 |
| R.F. Cardiovascular (No. %) | 16/36 (44.4%) | 10/68 (14.7%) | 0.001 |
| R.F. Obesity (No. %) | 8/36 (22.2%) | 11/68 (16.2%) | 0.453 |
| R.F. Diabetes (No. %) | 7/36 (19.4%) | 10/68 (14.7%) | 0.538 |
| R.F. Pulmonary (No. %) | 4/35 (11.1%) | 2/68 (2.9%) | 0.099 |
| R.F. Renal (No. %) | 3/36 (8.3%) | 2/68 (2.9%) | 0.962 |
| R.F. Pregnancy (No. %) | 1/36 (2.8%) | 1/68 (1.5%) | 0.652 |
| R.F. Neurologic (No. %) | 0/36 (0%) | 1/68 (1.5%) | 0.355 |
| R.F. Cancer (No. %) | 0/36 (0%) | 1/68 (1.5%) | 0.355 |
| R.F. Liver disease (No. %) | 0/36 (0%) | 0/68 (0%) | - |
| R.F. Immune (No. %) | 0/36 (0%) | 0/68 (0%) | - |

Notes: R.F.: risk factor (disease). S.D.: standard deviation. No.: number. %: percent.

When the data regarding vital and respiratory functions were evaluated, it was found that there were statistically significant differences between both groups regarding the respiratory / oxygen evaluation such as: respiratory rate, arterial oxygen saturation (SaO2), PaFi (relationship between PaO2 / inspired oxygen fraction), SaFi (ratio between SaO2 / inspired oxygen fraction) and ROX index (SaFi / respiratory rate per minute); All these variables indicated a greater severity of respiratory / oxygen compromise in the group that received HCQ compared to the group that received TDF (Table No. 2). In addition, when SaFi values were stratified into three groups: mild (SaFi between 325 and 450), moderate (SaFi between 200 and 325) and severe (SaFi between 75-200), it was found that this distribution was more uniform in the HCQ group (38.9%, 30.6% and 30.6% respectively) in contrast to the patients in the TDF group where according to SaFi 57.4% were mild, 33.8% moderate and 8.8% severe (p = o.o18).

**Table No. 2.** Tenofovir-DF versus Hydroxychloroquine: vital and respiratory functions.

| Variables | Hydroxychloroquine<br>36 patients | Tenofovir<br>68 patients | p |
| --- | --- | --- | --- |
| Heart rate (per minute) | 96 ± 15 | 92 ± 16 | 0.1280 |
| Breathing frequency (per minute) | 25 ± 5 | 23 ± 4 | 0.0301 |
| Systolic B.P. (mm Hg) | 120 ± 19 | 119 ± 14 | 0.4340 |
| Diastolic B.P.(mm Hg) | 73 ± 13 | 72 ± 8 | 0.3419 |
| Temperature (°C) | 37.4 ± 0.7 | 37.3 ± 0.9 | 0.1973 |
| SaO<sub>2</sub> | 88 ± 8 | 92 ± 3 | 0.0016 |
| SaFi/resp.rate (ROX) | 11.8 ± 7.4 | 14.8 ± 5.2 | 0.0099 |
| PaFi | 213 ± 81 | 270 ± 59 | 0.0065 |
| SaFi Total | 277 ± 117 | 336 ± 89 | 0.0023 |
| SaFi Grade mild: 325-450 | 14/36 (38.9%) | 39/68 (57.4%) | 0.015 |
| SaFi Grade moderate: 200-325 | 11/36 (30.6%) | 23/68 (33.8%) | 0.015 |
| SaFi Grade severe: 75-200 | 11/36 (30.6%) | 6/68 (8.8%) | 0.015 |
| CT scan: affected lung > 50% | 12/36 (33.3%) | 25/68 (36.8%) | 0.727 |

Notes: B.P.: blood pressure; mmHg: millimeters of mercury; °C: Celsius degrees; SaO<sub>2</sub>: arterial oxygen saturation; FiO<sub>2</sub>: inspired oxygen fraction of oxygen; ROX: ROX index (SaFi/breathing frequency per minute); PaFi: ratio between arterial oxygen partial pressure (PaO<sub>2</sub>) and FiO<sub>2</sub>; SaFi: ratio between arterial oxygen saturation (SaO<sub>2</sub>) and FiO<sub>2</sub>; CT scan: computed tomography, multislice spiral.

No statistically significant differences were found with heart rate, systolic or diastolic blood pressure and temperature (table No. 2). The frequency of pulmonary tomographic involvement by COVID-19 greater than 50% was also similar in both groups.

The results of the laboratory tests (Table No. 3) showed that there was no statistically significant difference between the HCQ group compared with the TDF group regarding: blood group A +, total leukocytes, filled or segmented neutrophils, monocytes, eosinophils, basophils, hemoglobin, hematocrit, platelet number, partial thromboplastin time (TTP), thrombin time (TT), globulin level, uric acid, cholesterol, total bilirubins, TGP (ALT), alkaline phosphatase, γ-glutamyl transpeptidase, creatine-Total kinase, creatine-kinase fraction MB, troponin, sodium, chlorine, and D-dimer levels.

**Table No. 3.** Tenofovir-DF versus Hydroxychloroquine: laboratory tests.

| Variables | Hydroxychloroquine<br>36 patients | Tenofovir<br>68 patients | p |
| --- | --- | --- | --- |
| Blood type A+ (No. %) | 1/11 (9.1%) | 2/26 (7.7%) | 0.888 |
| Leukocytes (cells/mm<sup>3</sup>) | 8663 ± 4055 | 8241 ± 4361 | 0.3162 |
| Neutrophil bands (cells/mm<sup>3</sup>) | 294 ± 449 | 577 ± 898 | 0.0751 |
| Neutrophils (cells/mm<sup>3</sup>) | 6754 ± 3651 | 5949 ± 3584 | 0.1405 |
| Lymphocytes (cells/mm<sup>3</sup>) | 1040 ± 567 | 1354 ± 595 | 0.0054 |
| Monocytes (cells/mm<sup>3</sup>) | 421 ± 279 | 490 ± 250 | 0.1011 |
| Eosinophils (cells/mm<sup>3</sup>) | 47 ± 51 | 45 ± 44 | 0.4233 |
| Basophils (cells/mm<sup>3</sup>) | 18 ± 25 | 23 ± 27 | 0.1850 |
| Hemoglobin (gr/100 ml) | 13.9 ± 1.9 | 14.3 ± 1.8 | 0.1550 |
| Hematocrit (%) | 40.9 ± 5.7 | 42.0 ± 4.9 | 0.1600 |
| Platelets (cells/mm<sup>3</sup>) | 260,408 ± 76,612 | 252,838 ± 122,529 | 0.3684 |
| P.T. (sec.) | 11.4 ± 1.2 | 10.8 ± 0.7 | 0.0013 |
| I.N.R. | 1.02 ± 0.13 | 0.96 ± 0.07 | 0.0034 |
| P.T.T. (sec.) | 33.8 ± 7.6 | 34.8 ± 5.1 | 0.0670 |
| T.T. (sec.) | 17.4 ± 1.0 | 16.0 ± 2.6 | 0.0700 |
| Glucose (mg/100 ml) | 124 ± 47 | 107 ± 41 | 0.0308 |
| Creatinin (mg/100 ml) | 1.21 ± 1.36 | 0.84 ± 0.89 | 0.0492 |
| Creatinin (mg/100 ml) no CKD | 0.88 ± 0.26 | 0.74 ± 0.18 | 0.0009 |
| Urea (mg/100 ml) | 46.7 ± 46.8 | 28.1 ± 10.0 | 0.0013 |
| Urea no CKD (mg/100 ml) | 37.0 ± 26.1 | 27.3 ± 8.0 | 0.0039 |
| Albumin (gr/100 ml) | 3.2 ± 0.6 | 3.5 ± 0.4 | 0.0028 |
| Globulins (gr/100 ml) | 3.1 ± 0.6 | 2.8 ± 0.5 | 0.0740 |
| Uric acid (mg/100 ml) | 2.6 ± 1.3 | 3.2 ± 2.0 | 0.2837 |
| Cholesterol (mg/100 ml) | 134.2 ± 33.2 | 135.5 ± 33.8 | 0.4467 |
| Triglycerides (mg/100 ml) | 195.1 ± 148.4 | 140.8 ± 53.0 | 0.0458 |
| Bilirubin, total (mg/100 ml) | 0.78 ± 0.48 | 0.65 ± 0.33 | 0.0628 |
| Bilirubin, direct (mg/100 ml) | 0.44 ± 0.35 | 0.32 ± 0.18 | 0.0180 |
| AST (TGO) (U/Litro) | 70.5 ± 48.4 | 48.8 ± 37.3 | 0.0072 |
| ALT (TGP) (U/Litro) | 73.7 ± 75.1 | 58.0 ± 45.0 | 0.0981 |
| Alkaline phosphatase (U/Litro) | 144.8 ± 98.8 | 120.3 ± 59.4 | 0.0681 |
| GGT (U/Litro) | 158.3 ± 149.3 | 145.2 ± 162.1 | 0.3749 |
| CPK (U/Litro) | 161.1 ± 267.8 | 120.2 ± 179.2 | 0.2316 |
| CPK-MB (U/Litro) | 17.8 ± 7.3 | 23.3 ± 14.6 | 0.0643 |
| Troponin (ng/ml) | 0.00945 ± 0.01451 | 0.00602 ± 0.00570 | 0.0716 |
| Na<sup>+</sup> (mEq/Litro) | 138.8 ± 5.1 | 139.2 ± 3.5 | 0.3253 |
| K<sup>+</sup> (mEq/Litro) | 4.1 ± 0.7 | 3.9 ± 0.5 | 0.0294 |
| Cl<sup>-</sup> (mEq/Litro) | 105.1 ± 7.8 | 103.4 ± 3.7 | 0.0959 |
| PCR (mg/100 ml) | 15.7 ± 12.0 | 11.2 ± 8.0 | 0.0144 |
| Ferritin (ng/ml) | 1198.6 ± 818.1 | 927.3 ± 671.4 | 0.0372 |
| Dímero-D (U/100 ml) | 2.86 ± 4.10 | 1.78 ± 5.82 | 0.1871 |
| Fibrinogen (mg/100 ml) | 718.8 ± 200.4 | 646.6 ± 130.1 | 0.0319 |
| DHL (U/Litro) | 389.6 ± 188.7 | 311.0 ± 89.7 | 0.0028 |

Notes: No.: number; %: percentage; mm<sup>3</sup>: cubic millimeter; gr.: grams; P.T.: prothrombin time; sec.: seconds; I.N.R.: international normalized ratio of P.T.; P.T.T.: partial thromboplastin time; T.T.: thrombin time; mg.: milligrams; C.K.D.: chronic kidney disease; T.G.O.: glutamic-oxalacetic transaminase; A.S.T.: aspartate aminotransferase; U.: units; T.G.P.: glutamic-pyruvic transaminase; A.L.T.: alanin aminotransferase; G.G.T.: γ-glutamyl transferase; C.P.K.: creatin phosphokinase; C.P.K.-M.B.: creatin phosphokinase MB fraction; ng: nanograms; Na<sup>+</sup>: sodium; mEq: milliequivalents; K<sup>+</sup>: potassium; Cl<sup>-</sup>: chlorine; P.C.R.: protein, c-reactive; D.H.L.: lactate-dehydrogenase.

Statistically significant differences were found, but without relevant clinical significance and also within the range of normal values for our laboratory a: prothrombin time (PT) and international normalized ratio (INR), triglycerides, direct bilirubin and serum potassium levels (K +), so these differences were not taken into account for the multivariate analysis.

There were clear statistical and clinical differences regarding higher glucose, creatinine, and urea levels in the HCQ group compared to the TDF group. In addition, a statistically significant decrease in albumin was found in the HCQ group compared to the TDF group (Table No. 3).

Likewise, total lymphocyte levels were significantly lower in the HCQ group. In addition, other markers of inflammation such as: C-reactive protein (CRP), ferritin, fibrinogen, and lactate dehydrogenase (DHL) levels were higher in HCQ patients. Regarding the D-dimer, no statistically significant difference was found between both groups (Table No. 3). Thus, the level of inflammatory markers estimated by a simplified cytokine storm score (STCQs, supplementary material) was high for 8/36 (22.2%), intermediate for 14/36 (38.9%) and low for 14/36 (38.9%) of patients in the HCQ group; while in the patients of the TDF group, the levels were high for 5/68 (7.4%), intermediate for 27/68 (39.7%) and low for 36/68 (52.9%) of the patients (p = o. o86).

Data on other therapies indicated to hospitalized patients with COVID-19 were also collected (Table No. 4). There was no significant difference in the use of corticosteroids (methyl-prednisolone), metamizole and enoxaparin between both groups. HCQ patients received lopinavir / ritonavir (LPV / r) more frequently (91.7% vs. 17.6%), azithromycin (58.3% vs. 16.2%), and atorvastatin (16.7% vs. 2.9%) compared to those in the TDF group: all these differences were statistically significant. Thus, it can be considered that the study actually compares the combination of HCQ plus LPV / r versus TDF alone; and to a lesser extent the same happened with the use of azithromycin, so it was not considered appropriate to include these two drugs in the multivariate analysis. Although there were significant differences with the use of atorvastatin, this was not considered for further analysis because of the following: there were only 8 patients (6 in the HCQ group and 2 in the TDF group) and of them 80% were in the ICU where it was usual practice to use said drug at that time. No patient received Tocilizumab.

**Table No. 4.** Tenofovir-DF versus Hydroxychloroquine: other therapies.

| **Variables** | **Hydroxychloroquine**<br>**36 patients** | **Tenofovir**<br>**68 patients** | **p** |
| --- | --- | --- | --- |
| Azithromycin (AZM) | 21/36 (58.3%) | 11/68 (16.2%) | 0.000 |
| Lopinavir/ritonavir (LPV/r) | 33/36 (91.7%) | 12/68 (17.6%) | 0.000 |
| Methyl-Prednisolone (MP) | 23/36 (63.9%) | 35/68 (51.5%) | 0.223 |
| Metamizole | 25/36 (69.4%) | 50/68 (73.5%) | 0.660 |
| Enoxaparin | 34/36 (94.4%) | 67/68 (98.5%) | 0.252 |
| Atorvastatin | 6/36 (16.7%) | 2/68 (2.9%) | 0.015 |
| Tocilizumab | 0/36 (0.0%) | 0/68 (0.0%) | - |

When the primary outcomes of the study were evaluated comparing the two groups, it was found that the LOS was 16.6 days for the HCQ group and 12.2 days for the TDF group; likewise, the need for intensive care (ICU) or need for mechanical ventilation (MV) was 61.1% and 11.8% respectively; and finally the mortality reached 50.0% and 8.8% respectively. All these results were statistically significant (Table No. 5) in the univariate analysis.

**Table No. 5.** Tenofovir-DF versus Hydroxychloroquine: primary outcomes.

| **Variables** |  | **Hydroxychloroquine**<br>**36 patients** | **Tenofovir**<br>**68 patients** | **p** |
| --- | --- | --- | --- | --- |
| Hospital stay |  | 16.6 ± 12.1 days | 12.2 ± 7.0 days | 0.0102 |
| SaFi Mild level: | 325-450 | 16.9 ± 14.7 days | 11.6 ± 7.7 days | 0.0452 |
| SaFi Moderate level: | 200-325 | 14.3 ± 8.3 days | 13.1 ± 5.8 days | 0.3158 |
| SaFi Severe level: | 75-200 | 18.4 ± 12.3 days | 12.3 ± 6.9 days | 0.1451 |
| Need for UCI/VM |  | 22/36 (61.1%) | 8/68 (11.8%) | 0.000 |
| SaFi Mild level: | 325-450 | 7/14 (50.0%) | 3/39(7.7%) | 0.001 |
| SaFi Moderate level: | 200-325 | 5/11 (45.4%) | 4/23 (17.4%) | 0.089 |
| SaFi Severe level: | 75-200 | 10/11 (90.9%) | 1/6 (16.7%) | 0.002 |
| Deceased |  | 18/36 (50.0%) | 6/68 (8.8%) | 0.000 |
| SaFi Mild level: | 325-450 | 6/14 (42.9%) | 1/39 (2.6%) | 0.000 |
| SaFi Moderate level : | 200-325 | 4/11 (36.4%) | 3/23 (13.0%) | 0.126 |
| SaFi Severe level: | 75-200 | 8/11 (72.7%) | 2/6 (33.3%) | 0.113 |

Notes: SaFi: SaO<sub>2</sub>/FiO<sub>2</sub>; V.M.: ventilation, mechanic; U.C.I.: intensive care unit.

Multivariate analyzes were performed to adjust the outcomes to the differences in the two groups that had been demonstrated in the univariate analysis: demographic factors: male sex and cardiovascular risk factor (Table 1): the relevant tests of vital functions and respiratory involvement: SaFi (Table No. 2) and laboratory tests for markers of chronic disease: glucose, creatinine and albumin; and inflammation (Table No. 3). For inflammation markers, an inflammation or “cytokine storm” score was used, which is attached to the supplementary material. Furthermore, for the purpose of tightening the analysis, four patients (one from the HCQ group and 3 from the TDF group) who were the only ones who did not receive oxygen during their hospitalization are not considered.

In the patients who did not die, the LOS was 17.7 ± 11.8 days for the HCQ group compared to 11.5 ± 6.4 days for the TDF group (p = o.oo25). The patients with HCQ who died had a LOS of 15.4 ± 12.6 days and in those with TDF it was 18.7 ± 9.7 days (p = o.7130). Table No. 6 shows the results of the multiple linear regression model for the LOS variable in all patients. There are statistically significant differences only for TDF use and sex. When the stays were stratified by the severity of the respiratory compromise defined by the SaFi levels (Table No. 5), it was found that in mild cases (SaFi from 325-450) the stay was 16.9 ± 14.7 for HCQ vs. 11.6 ± 7.7 days for TDF (p = o.o452), in the moderate ones (SaFi of 200-325) it was 14.3 ± 8.3 for HCQ vs. 13.1 ± 5.8 days for TDF (p = o.3158) and in the severe ones (SaFi between 75-200) it was 18.4 ± 12.3 for HCQ vs. 12.3 ± 6.9 days for TDF (p = o.1451).

**Table No. 6.** Tenofovir-DF versus Hydroxychloroquine: multiple regression model for hospital stay.

| Variable | Coefficient | p | 95% Confidence Interval |  |
| --- | --- | --- | --- | --- |
| Tenofovir | -6.104518 | 0.042 | -11.96887 | -.2401695 |
| Male sexo | -5.741626 | 0.050 | -11.49024 | .0069907 |
| R.F. CDV | -5.85061 | 0.100 | -12.85043 | 1.149206 |
| SaFi | -.0029125 | 0.841 | -.0319378 | .0261127 |
| Glucose | .0236312 | 0.402 | -.0324615 | .0797239 |
| Creatinin | .1644922 | 0.871 | -1.855607 | 2.184591 |
| Albumin | -2.027808 | 0.548 | -8.752981 | 4.697364 |
| STCQs | 1.248916 | 0.547 | -2.883485 | 5.381316 |
| -constant: | 25.22768 | 0.064 | -1.571162 | 52.02653 |

Notes: R.F.: risk factor; C.D.V.: cardiovascular; SaFi: arterial oxygen saturation (SaO<sub>2</sub>) divided by the inspired oxygen fraction (FiO<sub>2</sub>); S.T.C.Q.s.: simplified cytokine storm score.

In the same way, the results of the multivariate analysis of the need for intensive care (ICU) or mechanical ventilation (MV) are presented in Table No. 7. There were statistically significant differences only for the use of TDF compared to the use of HCQ and with serum albumin levels. When stratifying the need for ICU / MV by the degree of involvement defined by the SaFi (Table No. 5), it was found that in mild cases (SaFi from 325-450) the need for ICU / MV reached 50.0% for HCQ vs. 7.7% for TDF (p = o.oo1), in the moderate ones (SaFi of 200-325) it reached 45.4% for HCQ vs. 17.4% for TDF (p = o.o89) and in the severe (SaFi between 75-200) it reached 90.9% for HCQ vs. 16.7% for TDF (p = o.oo2).

The multivariate Cox proportional hazards model for mortality is shown in Table No. 8. Here it was found that there were statistically significant differences for: the use of TDF (HR 0.163 p = o.o45) and serum albumin levels (HR 0.141 p = o.o18). In this model, no difference was found for: male sex, presence of cardiovascular risk factor (F.R. CDV), SaFi, glucose levels, creatinine or simplified cytokine storm score (STCQs).

**Table No. 7.** Tenofovir-DF versus Hydroxychloroquine: logistic regression model for the need for UCI/VM.

| Variable | O.R. | p | Intervalo de Confianza al 95% |  |
| --- | --- | --- | --- | --- |
| Tenofovir | .1511135 | 0.022 | .0298717 | .7644461 |
| Male sex | .4707254 | 0.395 | .0829883 | 2.670044 |
| R.F. CDV | .9503795 | 0.958 | .1452307 | 6.219217 |
| SaFi | 1.000384 | 0.931 | .9917411 | 1.009102 |
| Glucose | .9988181 | 0.871 | .9846474 | 1.013193 |
| Creatinin | 1.014047 | 0.975 | .423668 | 2.427115 |
| Albumin | .0389615 | 0.010 | .0032763 | .4633254 |
| STCQs | 1.624817 | 0.399 | .5256538 | 5.022371 |

Notes: UCI: intensive care unit; VM: mechanic ventilation; R.F: risk factor; C.D.V.: cardiovascular; SaFi: arterial oxygen saturation (SaO<sub>2</sub>) divide by the inspired oxygen fraction (FiO<sub>2</sub>); S.T.C.Q.s.: simplified cytokine storm score.

**Table No. 8.** Tenofovir-DF versus Hydroxychloroquine: Cox proportional risk model for mortality.

| Variable | Hazard ratio | p | 95% Confidence Interval |  |
| --- | --- | --- | --- | --- |
| Tenofovir | .1628898 | 0.041 | .0275778 | .9621179 |
| Male sex | .7198004 | 0.714 | .1237367 | 4.187219 |
| R.F. CDV | 2.158883 | 0.207 | .6532757 | 7.134467 |
| SaFi | 1.00241 | 0.451 | .9961562 | 1.008704 |
| Glucose | 1.001362 | 0.822 | .9895546 | 1.01331 |
| Creatinin | .7355847 | 0.074 | .5250609 | 1.030518 |
| Albumin | .1411998 | 0.018 | .0279286 | .7138683 |
| STCQs | 2.367026 | 0.083 | .8926457 | 6.276636 |

Notes: R.F: risk factor; C.D.V.: cardiovascular; SaFi: arterial oxygen saturation (SaO<sub>2</sub>) divided by the inspired oxygen fraction (FiO<sub>2</sub>); S.T.C.Q.s.: simplified cytokine storm score.

In Graph No. 1 the Kaplan-Meier survival curves for mortality due to the use of Hydroxychloroquine (HCQ) or tenofovir (TDF) are shown, the 95% confidence intervals are also shown in the same graph. In the same way, the Kaplan-Meier survival curves stratified by SaFi levels with 95% confidence intervals are presented: in Graph No. 2, patients with mild SaFi (325-400); in Graph No. 3 the patients with moderate SaFi (200-325) and in Graph No. 3 the patients with severe SaFi levels (75-200). Although there is a trend towards better survival in the TDF group compared to HCQ in all categories, the results were statistically significant only for the global ones (p = o.ooo3) and for the level of mild affectation according to the SaFi level (325-400) (p = o.ooo1) and not for moderate or severe levels of SaFi (p = o.o852 and p = o.2336 respectively) according to the stratified log-rank test for the equality of functions test of survival. One patient in the HCQ group died at 61 days and one in the TDF group at 35 days, so these two patients do not appear in the Kaplan-Meier survival curves of 30 days.

**Graphic No. 1.**
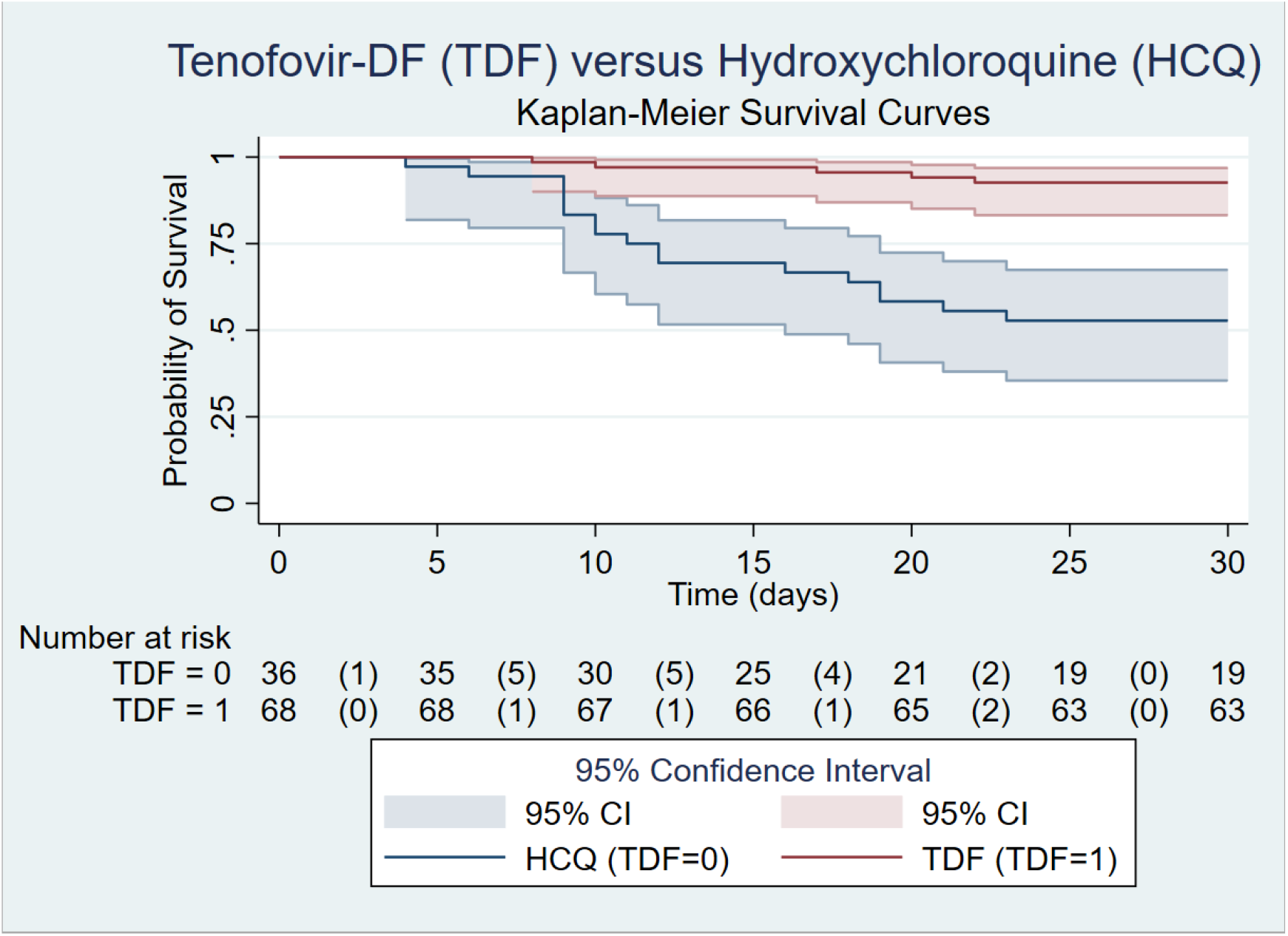
Tenofovir-DF versus Hydroxychloroquine: Global Kaplan-Meier survival curves with 95% confidence interval.

**Graphic No. 2.**
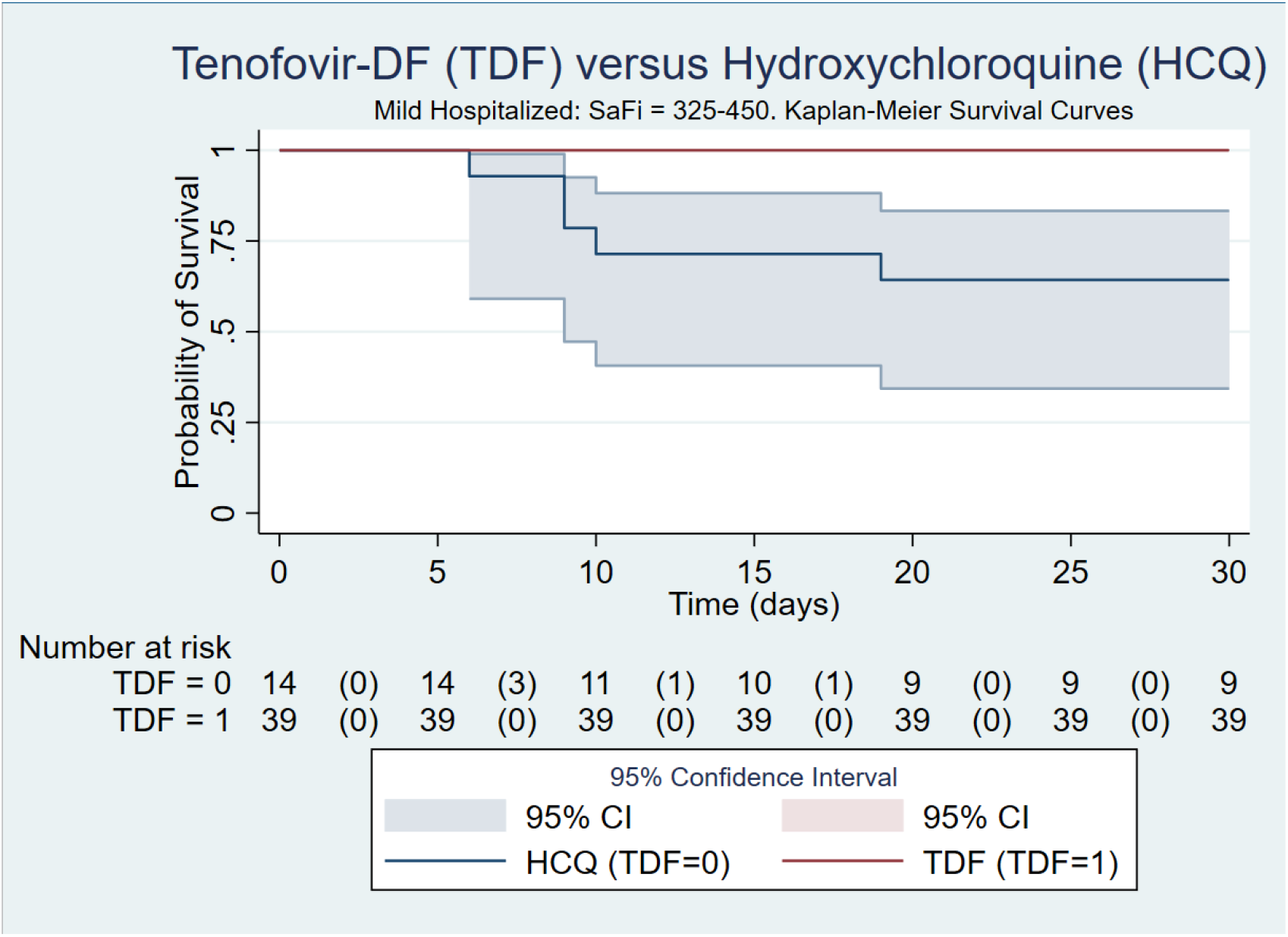
Tenofovir-DF versus Hydroxychloroquine: Kaplan-Meier survival curves for hospitalized with mild illness: patients with SaFi levels between 325-450 (with 95% confidence interval).

**Graphic No. 3.**
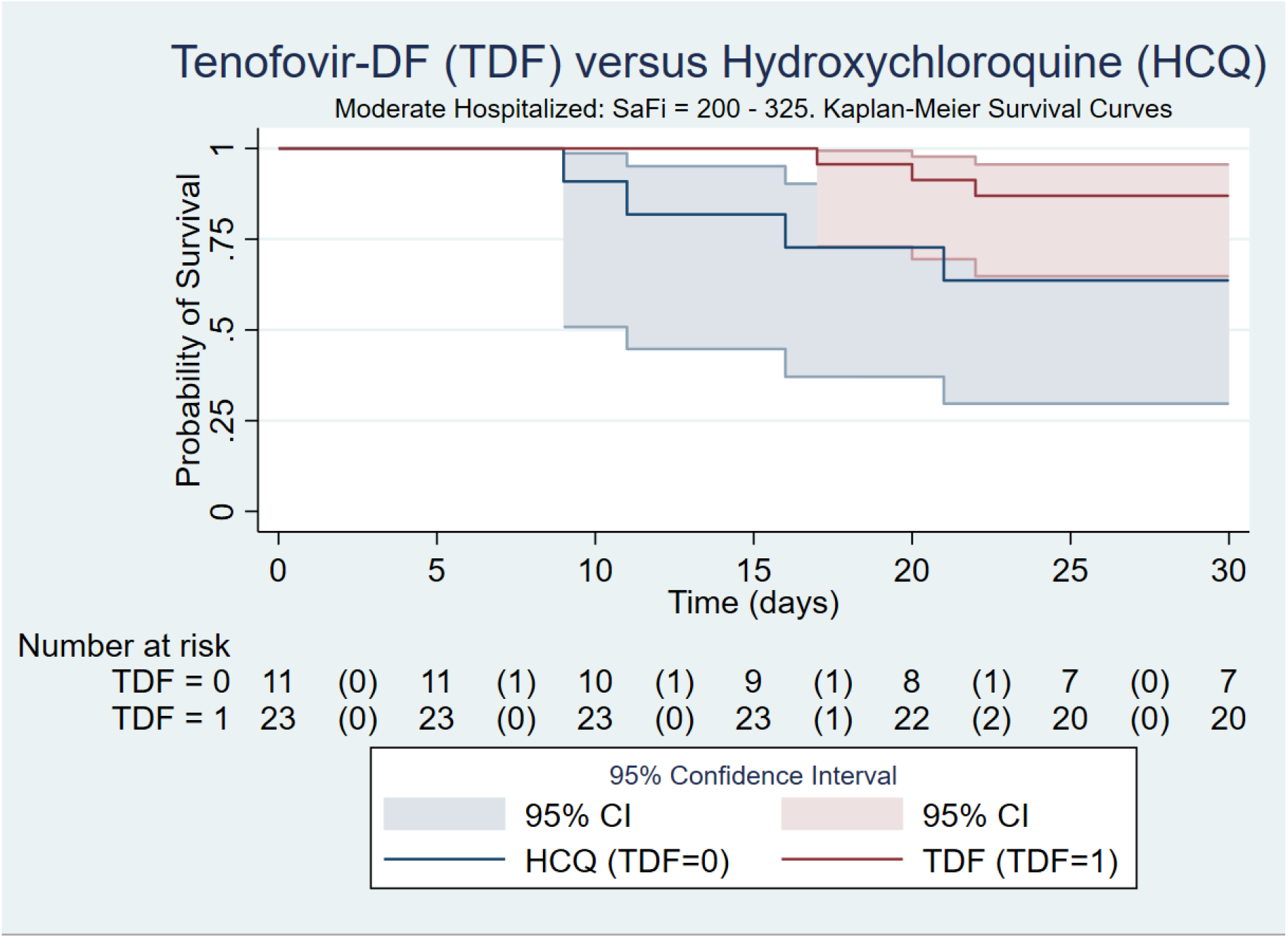
Tenofovir-DF versus Hydroxychloroquine: Kaplan-Meier survival curves for hospitalized with moderate illness: patients with SaFi levels between 200-325 (with 95% confidence interval).

**Graphic No. 4.**
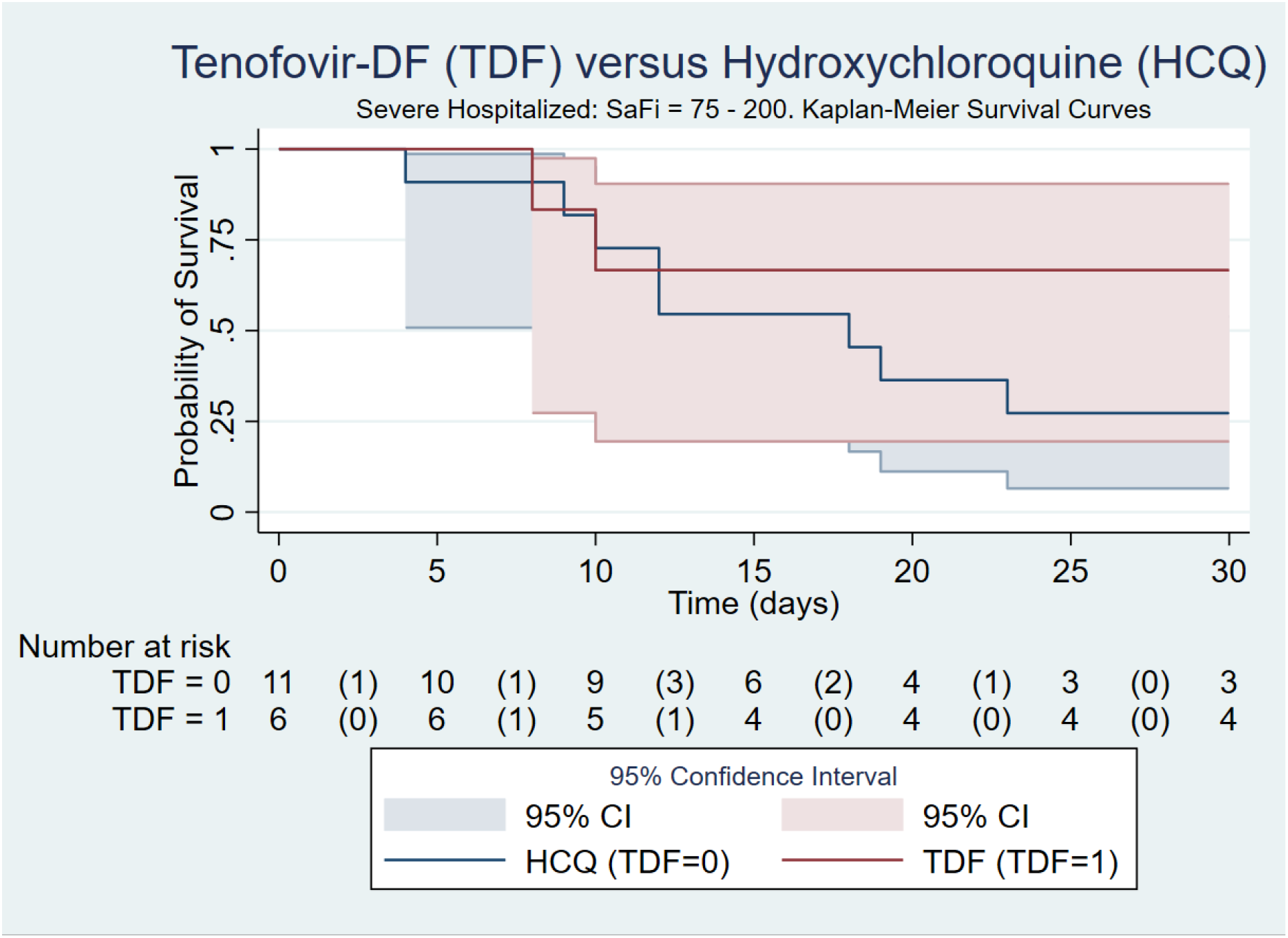
Tenofovir-DF versus Hydroxychloroquine: Kaplan-Meier survival curves for hospitalized with severe illness: patients with SaFi levels between 75-200 (with 95% confidence interval).

The overall mortality of the HCQ group vs. the TDF group was 50.0% vs. 8.8% (Table No. 5). As there were differences in the severity of the disease upon admission to the hospital, the two groups were stratified according to the SaFi level (Table No. 5); thus, the mortality values were: Mild SaFi group 42.9% for HCQ vs. 2.6% for TDF (p = o.ooo), in the moderate SaFi group 36.4% for HCQ vs. 13.0% for TDF (p = 0.126); and in the severe SaFi group 72.7% for HCQ vs. 33.3% for TDF (p = 0.113), using the chi2 test.

In Graph No. 5 the patients who died are stratified by their inflammatory score or “cytokine storm” (STCQs, supplementary material) upon admission to the hospital: it was found that in the HCQ group, for each point the mortality it increased consistently in an almost linear way: 33%, 27%, 44%, 60%, 86% and 100% (p = o.o21); On the other hand, in the TDF group the values were lower and the line was flatter and with less slope (p = o.577). In Graph No. 6 the mortality data are presented according to the highest inflammatory score (STCQs) during the first 7 days of hospitalization. In this case if a linear relationship can be seen with respect to higher mortality at higher STCQs both for the HCQ (p = o.oo8) and for the TDF (p = o.oo9).

**Graphic No. 5.**
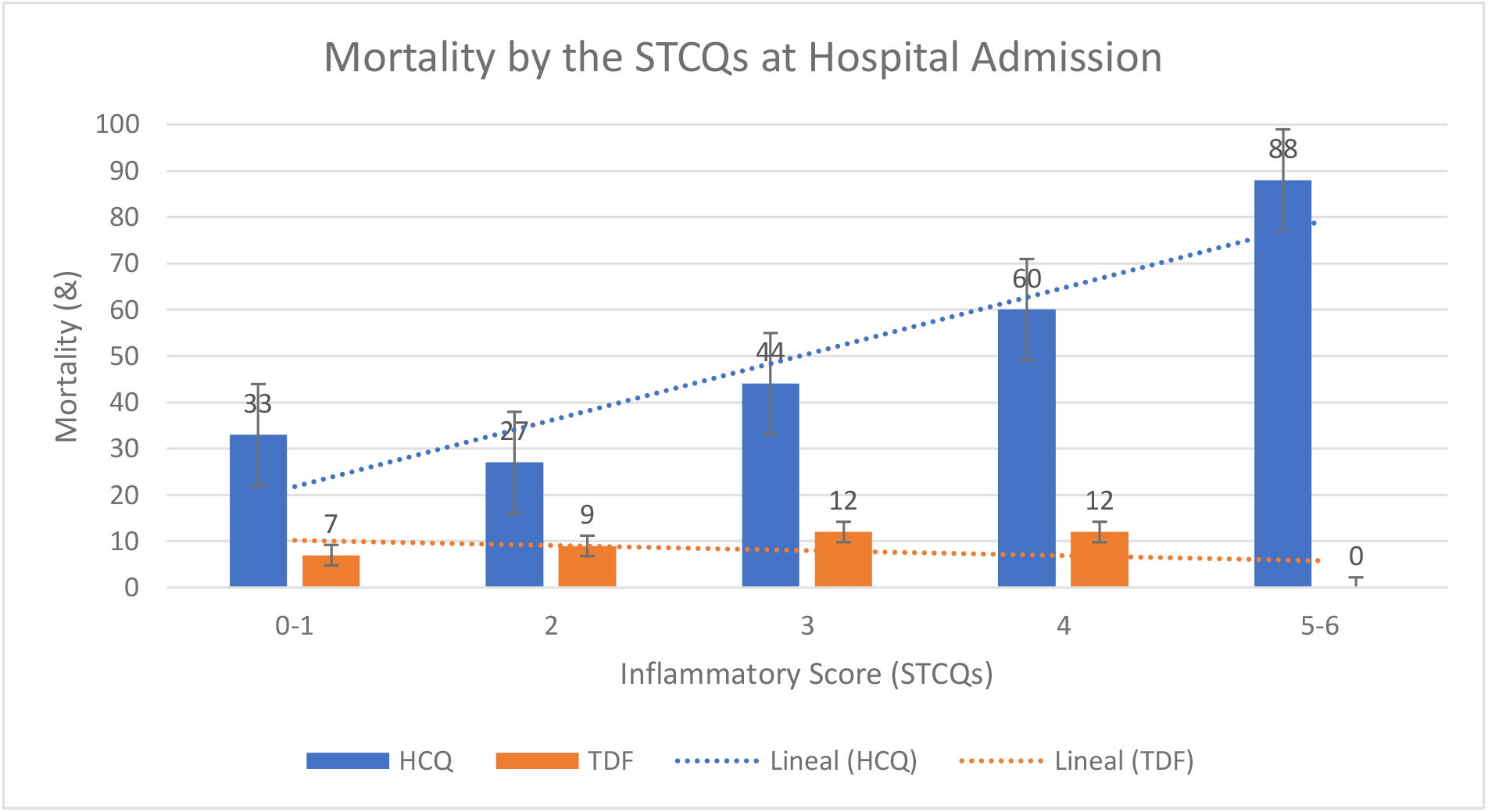
Tenofovir-DF versus Hydroxychloroquine: TDF compared with HCQ: mortality due to the inflammatory score or “cytokine storm” (STCQs) on admission to hospital. Notes: The relationship between mortality and the STCQs at hospital admission for HCQ was significant (p=o.o21) but not for TDF (p=o.577).

**Graphic No. 6.**
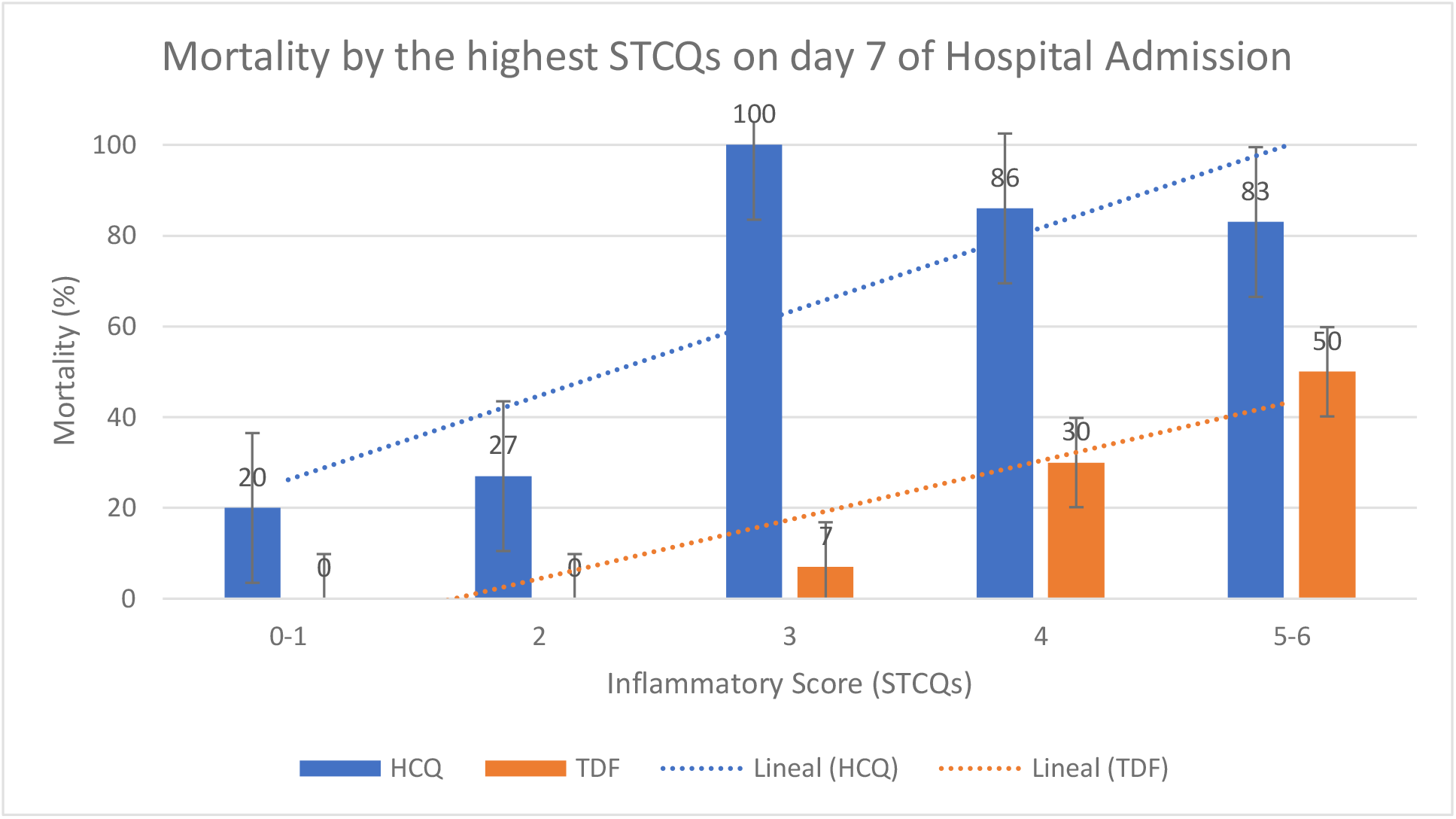
Tenofovir-DF versus Hydroxychloroquine: mortality due to the highest inflammatory score or “cytokine storm” (STCQs) within 7 days after admission to the hospital. Notes: The relationship between mortality and increased STCQs within 7 days of hospital admission was significant for both HCQ (p=o.oo8) and TDF (p=o.oo9).

Per judgment of the treating physicians regarding the study drugs, 13 adverse events were recorded in 36 patients in the HCQ group (36%): 3 of them were acute diarrheal disease associated with the use of LPV / r, that is, 3/45 (6.7%). The 10 adverse effects associated with the use of HCQ were cardiac and led to discontinuation of the drug in all cases; thus, its frequency reached 10/36 (27.8%) and were the following: 5 extreme prolongations of the QTc interval (corrected), 2 cases with prolonged QTc and bradycardia, 2 cases with isolated bradycardia and 1 case of complex tachycardia that required cardioversion electric four times. No adverse effects associated with the use of TDF were reported.

Furthermore, since it is an observational study and with evident baseline differences in the two groups, models were developed (Table No. 9) for estimating the effects of TDF treatment (only counting patients who required oxygen) by regression adjustment. for the primary outcomes of the study: hospital stay, need for ICU (intensive care unit) and / or MV (mechanical ventilation) and mortality. All of them were adjusted for sex, cardiovascular risk factor, SaFi levels, glucose, creatinine, albumin, and simplified cytokine storm score (STCQs). In the three outcomes, the difference in the use of TDF compared to HCQ was statistically significant: the use of TDF reduced the hospital stay by an average of 6.4 days, decreased the need for ICU / MV by about 42%, and decreased mortality in about 35% (Table No. 9).

**Table No. 9.** Tenofovir-DF versus Hydroxychloroquine: model for estimating the effects of TDF treatment by regression adjustment for the primary outcomes.

| Variable | Coefficiente<br>(with TDF) | Coefficient<br>(w/o TDF) | p | 95% Confidence Interval |  |
| --- | --- | --- | --- | --- | --- |
| Hospital stay | -6.380131 | 17.53772 | 0.036 | -12.34486 | -.415402 |
| UCI/VM | -.4174426 | .533719 | 0.000 | -.6371957 | -.1976896 |
| Mortality | -.3521473 | .4185657 | 0.001 | -.5647387 | -.1395558 |

Notes: U.C.I.: intensive care unit; V.M.: mechanic ventilation.

## Discussion

At the beginning of the outbreak in Arequipa (March-April 2020), Hydroxychloroquine with or without lopinavir / ritonavir or azithromycin was used as a standard treatment and according to the best evidence at that time as a therapy against SARS-CoV-2 according to national guidelines [1] and the medical literature available [2-16]. However, monitoring the evolution of patients with these treatments showed us that what we saw was simply the natural history of the disease. In addition, some patients had side effects of the use of these drugs (QT prolongation, arrhythmias with Hydroxychloroquine or diarrhea with lopinavir / ritonavir). As it was impossible then and now to procure remdesivir in our workplace, it was important to find other alternatives within our reach.

Tenofovir-DF (disoproxil fumarate) (TDF) is an anti-retroviral agent with extensive experience in the world that is used in the treatment of HIV / AIDS infection. This nucleoside reverse transcriptase inhibitor has a good safety profile, its most important problems being the decrease in bone mineral density and kidney damage, including a Fanconi syndrome, induced the most severe effect and that is associated with its prolonged use; when TDF-induced kidney injury is detected early, it is reversible [1-8]. Other advantages of TDF are its low cost, the possibility of oral administration and its easy accessibility.

Both remdesivir and tenofovir (TDF) have been shown to bind strongly to SARS-CoV-2-dependent RNA polymerase [1, 2]. In a study in ferrets, the combination of TDF-ETC (emtricitabine) produced a decrease in viral titers [3]. In addition, recently a couple of studies of HIV positive patients on anti-retroviral therapy with TDF / FTC showed that they have a lower risk of COVID-19 and severe disease and admission to ICU [4, 5], this beneficial effect of tenofovir it was only apparent in tenofovir-disoproxil fumarate (TDF) users and not in tenofovir-alafenamide (TAF) users [6, 7]. This difference could be due to the fact that TAF is a more recently developed tenofovir prodrug that produces an intracellular concentration in peripheral mononuclear cells four times higher but a 90% lower plasma concentration of tenofovir than TDF [8-10]. If we consider COVID-19 as a systemic disease that strongly affects the endothelium [11-17] and that the viral load of SARS-Cov-2 in the blood correlates with the severity of the disease [18-22], the difference effectiveness of TDF compared to TAF could be explained.

Connexins and pannexins are two families of proteins that form channels that allow the passage of small signaling molecules and that have recently been shown to have multiple functions [1]. In particular, pannexin-1 has been shown to play an important role in endothelial inflammation and permeability, in the recruitment and activation of leukocytes, in the activation of platelets and other immune cells [1-6]; thus, its activity could explain, at least in part, the massive lung inflammation that can develop in severe COVID-19 [5]. TDF, but not TAF, has been shown to inhibit pannexin-1 [5, 7], which may explain why TDF appears to work and TAF does not in SARS-Cov-2 disease.

Unfortunately, as is often the case in observational studies, the presence of biases is practically unavoidable: thus, the patients in the HCQ group had a higher proportion of more severe COVID-19 disease, so the results had to be adjusted by statistical methods. Although the trend in outcomes favors TDF at all levels of severity, this difference was clearer in the group with less involvement, which is consistent with the general agreement that antiviral treatment is more effective in the first days of illness before full-scale inflammatory phase develops [1-3].

Likewise, it cannot be ruled out that perhaps a part of the benefit of TDF was due not only to the lack of efficacy of HCQ, but also to a deleterious effect of the drug itself [1-4]. The results presented in Graph No. 5 and No. 6 seem to clearly demonstrate the lack of clinical activity of the anti-inflammatory properties proposed for HCQ in COVID-19: there is a linear relationship from higher mortality to higher STCQs, which seems to indicate that the use of HCQ would not change the natural history of the disease; on the other hand, the inverse occurs if it is outlined for the TDF.

## In conclusion

in this observational study of the use of TDF compared to HCQ, despite recruiting only 104 patients, differences were shown in favor of the use of TDF in the three primary outcomes of the study: about 6 days less LOS, a decrease about 5 times the need for ICU / MV and 8 times less mortality. These differences were statistically significant overall and more marked for COVID-19 patients with SaFi levels greater than 325. There was a sustained trend for this to occur at all levels of severity defined by the SaFi level, but without statistical significance (except regarding the need for ICU / MV in the severe SaFi group), probably due to the small number of patients.

Despite the favorable results of TDF in this study, prompt randomized clinical trials are essential to confirm these findings.

## Data Availability

The data on which this study is based are available at
https://drive.google.com/file/d/1JADVw7uUkbbZLE7VphnvvkvzlGV1WdRr/view?usp=sharing

## Authors’ Contributions

MCG has participated in the design, data collection, analysis and interpretation of data and preparation of the article. RR and JS have participated in the design, data collection, review of the article and approval of the final version. NA, EB and JV have participated in the design and data collection. JC, YF, PL and CC have participated in the data collection.

## Ethics Review/Human Subjects Review

Board Status: Approved. Approval Number: NIT:2803-2021-07. Board Name: “Comité Institucional de Ética en Investigación de la Red Asistencial Arequipa (RAR) - EsSalud”. Board Affiliation: Instituto Nacional de Salud – Peru, RCEI-83 / IETSI.

## ClincalTrials.gov Identifier

NCT04812496.

## Acknowledgments

A special thanks to the Epidemiology team of the CASE-EsSalud National Hospital, made up of Amparo Villanueva, Monica Alvarez, Paola Cucho and Leit Huamani for their help in capturing, detecting and monitoring cases and confirmatory tests of SARS-Cov-2. Thanks also to Angela Eguren for the final review of the English translation.

## SUPPLEMENTS

Hospital Nacional CASE-EsSalud: Normal laboratory values.

Hospital Nacional CASE-EsSalud: Normal laboratory values.
| Variables | Normal Range | Units |
| --- | --- | --- |
| Blood Group A+ | - | - |
| Leukocytes | 5,000- 10,000 | cells/mm<sup>3</sup> |
| Neutrophils bands | 0-5 | % |
| Neutrophils total | 2.0-7.5 | % |
| Lymphocytes | 1-4 | % |
| Monocytes | 0.2-1 | % |
| Eosinophils | 0-0.5 | % |
| Basophils | 0-0.2 | % |
| Hemoglobin | 14-17 | gr/dl |
| Hematocrit | 45-52 | % |
| Platelets | 150,000-450,000 | cells/mm<sup>3</sup> |
| P.T. | 9.8-13.5 | sec. |
| I.N.R. | 0.9-1.1 | - |
| P.T.T. | 23-35 | sec. |
| T.T. | 16-21 | sec. |
| Glucose | 60-110 | mg/dl |
| Creatinin | 0.7-1.2 | mg/dl |
| Urea | 10-50 | mg/dl |
| Albumin | 3.5-5.2 | gr/dl |
| Globulins | 2.0-3.8 | gr/dl |
| Uric acid | 2.4-7.0 | mg/dl |
| Cholesterol | 150-200 | mg/dl |
| Tryglicerides | 30-200 | mg/dl |
| Bilirubin, total | 0.1-1.0 | mg/dl |
| Bilirubina, direct | 0.0-0.3 | mg/dl |
| TGO (AST) | 0-40 | U/L |
| TGP (ALT) | 0-41 | U/L |
| Alkalyn phosphatase | 35-105 | U/L |
| GGT | 5-36 | U/L |
| CPK | 39-308 | U/L |
| CPK-MB | 7-25 | U/L |
| Troponin | 0.00-0.03 | ng/ml |
| Na<sup>+</sup> | 135-145 | mEq/L |
| K<sup>+</sup> | 3.5-5.2 | mEq/l |
| Cl<sup>-</sup> | 95-105 | mEq/L |
| PCR | 0.0-0.5 | mg/dl |
| Ferritin | 20-200 | ng/ml |
| Dímero-D | 0.0-0.5 | U/dl |
| Fibrinogen | 200-400 | mg/dl |
| DHL | 135-225 | U/dl |

Abbreviations: P.T.: prothrombin time; I.N.R.: international normalized ratio of the P.T.; P.T.T.: partial thromboplastin time; T.T.: thrombin time; T.G.O.: transaminase, glutamic-oxalacetic; A.S.T.: aspartate aminotransferase; T.G.P.: transaminase, glutamic-piruvic; A.L.T.: alanin aminotransferasa; G.G.T.: γ-glutamyl transferase; C.P.K.: creatin phosphokinase; C.P.K.-M.B.: creatin phosphokinase MB-fraction; Na<sup>+</sup>: sodium; K<sup>+</sup>: potassium; Cl<sup>-</sup>: chlorine; P.C.R.: c-reactive protein; D.H.L.: lactate-dehydrogenase. mm<sup>3</sup>: cubic millimeter; gr.: grams; dl: deciliter; sec.: seconds; mg.: milligrams; U.: international units; $\mu$ g: micrograms; ng: nanograms; mEq: milliequivalents.

Inflammatory Score / “Cytokine Storm (TCQs)”.

**Inflammatory Score / “Cytokine Storm (TCQs)”.**
|  |  | POINTS | SCORE |
| --- | --- | --- | --- |
| **1** | **LYMPHOPENIA** |  |  |
|  | > 500 | 0 |  |
|  | < 500 | 1 | 1 |
| **2** | **PCR** |  |  |
|  | < 10 | 0 |  |
|  | > 10 | 1 | 1 |
| **3** | **FERRITIN** |  |  |
|  | < 1000 | 0 |  |
|  | > 1000 | 1 | 1 |
| **4** | **FIBRINOGEN** |  |  |
|  | < 600 | 0 |  |
|  | > 600 | 1 | 1 |
| **5** | **LACTATE DEHYDROGENASE (DHL)** |  |  |
|  | < 400 | 0 |  |
|  | > 400 | 1 | 1 |
| **6** | **TGO (AST)** |  |  |
|  | < 100 | 0 |  |
|  | > 100 | 1 | 1 |
| **TOTAL =** |  |  | **6** |

| TCQs PROBABILITY |  |
| --- | --- |
| SCORE MAXIMUM = 6 |  |
| 1 - 2 | LOW |
| 3 - 4 | MODERATE |
| 5 - 6 | HIGH |

